# Avoiding bias in Mendelian randomization when stratifying on a collider

**DOI:** 10.1101/2021.08.17.21262178

**Authors:** Claudia Coscia, Dipender Gill, Raquel Benítez, Teresa Pérez, Núria Malats, Stephen Burgess

## Abstract

**Background:** Mendelian randomization (MR) uses genetic variants as instrumental variables to investigate the causal effect of a risk factor on an outcome. A collider is a variable influenced by two or more other variables. Naive calculation of MR estimates in strata of the population defined by a variable affected by the risk factor can result in collider bias.

**Methods:** We propose an approach that allows MR estimation in strata of the population while avoiding collider bias. This approach constructs a new variable, the residual collider, as the residual from regression of the collider on the genetic instrument, and then calculates causal estimates in strata defined by quantiles of the residual collider. Estimates stratified on the residual collider will typically have an equivalent interpretation to estimates stratified on the collider, but they are not subject to collider bias. We apply the approach in several simulation scenarios considering different characteristics of the collider variable and strengths of the instrument. We then apply the proposed approach to investigate the causal effect of smoking on bladder cancer in strata of the population defined by bodyweight.

**Results:** The new approach generated unbiased estimates in all the simulation settings. In the applied example, we observed a trend in the stratum-specific MR estimates at different bodyweight levels that suggested stronger effects of smoking on bladder cancer among individuals with lower bodyweight.

**Conclusions:** The proposed approach can be used to perform MR studying heterogeneity among subgroups of the population while avoiding collider bias.

## Introduction

Mendelian randomization (MR) is the use of genetic variants as instrumental variables to assess the causal relationship between a risk factor and an outcome [1,2]. A valid instrumental variable (IV), or genetic instrument, must meet the following assumptions [3]: IV1, the instrument is associated with the risk factor; IV2, the instrument cannot affect the outcome directly, only potentially indirectly via the risk factor; and IV3, the instrument is not associated with any measured or unmeasured confounders (Figure 1A). If these assumptions are satisfied, an association of the instrument with the outcome is indicative of a causal effect of the risk factor on the outcome [1,4]. However, if either the IV2 or IV3 assumption are not satisfied, then the instrument could be associated with the outcome in the absence of a causal effect of the risk factor. However, only the IV1 assumption can be verified based on measured data [5].

**Figure 1.**
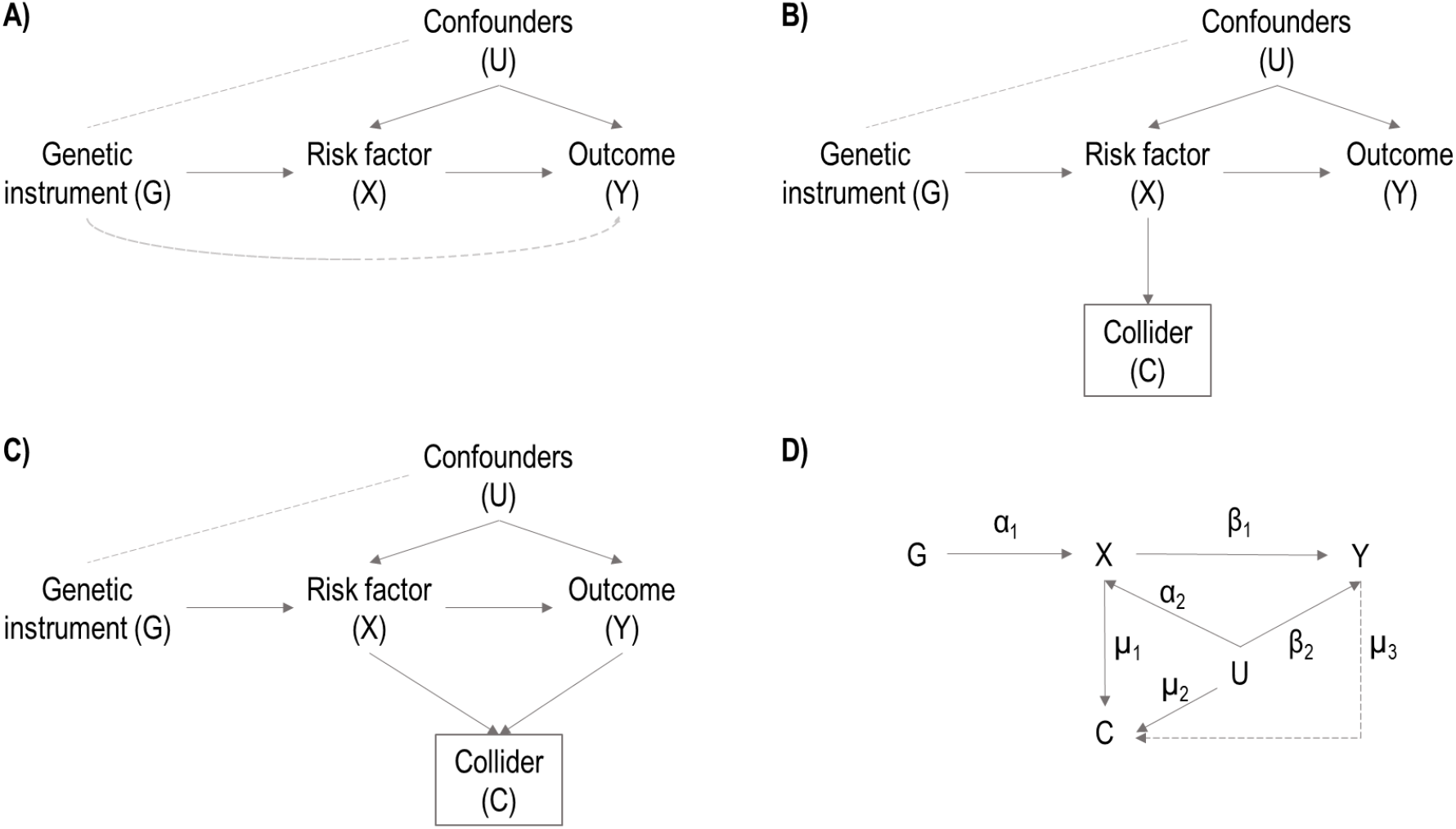
Directed Acyclic Graphs (DAGs) illustrating relationships between the variables. A) Mendelian Randomization causal diagram with the instrumental variable assumptions. The dashed lines between G and Y and between G and U, represent violations of the IV2 and IV3 assumptions respectively. B) DAG considering a collider variable C, being a common child of genetic instrument G and confounders U. When conditioning on C (indicated by the square box on C), G and U become correlated (dashed line between G and U) and a violation of the IV3 assumption occurs. C) DAG considering a collider variable C, being a common child of risk factor X and outcome Y. D) DAG illustrating the variables and parameters used for the simulation study. Dash line from Y to C correspond to simulation scenarios B1 to B3

Collider bias can occur when conditioning on a collider, defined as a variable that is a common effect of two or more variables [6,7]. The presence of collider can be recognized in a causal diagram when there are two arrows pointing at the same variable; the node at which the arrowheads “collide” together is a collider. For example, in the standard MR diagram, the risk factor is a collider as it is affected by both the instrument and the confounders. Moreover, any variable that is a causal descendent of collider is also affected by the same variables and so is itself a collider; hence in MR any variable influenced by the risk factor is a collider (Figure 1B). Even if the variables influencing a collider are independent, they will typically become dependent when conditioning on the collider. Hence conditioning on a variable affected by the risk factor will typically generate a conditional association between the instrument and the confounders, violating the IV3 assumption, and biasing Mendelian randomization estimates of the risk factor on the outcome.

Selection bias is a form of collider bias that occurs when selection of individuals into a dataset is dependent on a collider. For example, when disease progression is considered as an outcome, only patients who have already developed the disease would be recruited into the study [6]. If risk of developing the disease is influenced by the risk factor, then it is a collider when considering disease progression as the outcome, and selection of the study sample would result in collider bias. Several papers related to selection bias in the context of IV analysis and MR have been already published [8–10].

Collider bias could also occur when stratifying the population based on a collider. As an example, we consider investigating the causal effect of the risk factor on the outcome for individuals with specific levels of a stratifying variable. Stratification is important for identifying whether there are subgroups of the population for which causal effects of the risk factor are different, and so the outcome would be affected more strongly by an intervention on the risk factor. However, if the stratifying variable is a collider, an association between the instrument and the outcome in strata of the population could arise due to collider bias, invalidating the results. In particular, collider bias could affect some estimates more than others, leading to heterogeneity in the stratum-specific causal estimates even if the true causal effect is the same across strata.

The aim of this paper is to present an MR approach that obtains estimates in strata of the population that do not suffer from collider bias. The structure of this paper is as follows: first, we demonstrate the bias that arises from conditioning on a collider; second, we propose an approach to calculate MR estimates in strata of the population and evaluate heterogeneity between estimates in the different strata; third, we illustrate this new technique in simulation studies and an applied example using the UK Biobank resource; and finally, we discuss the interpretation of estimates and limitations of the approach.

## Methods

### Illustration of collider bias

The simplest MR method to estimate the causal effect of a risk factor *X* on outcome *Y* with a genetic instrument *G* is the ratio method [2]. With a single instrument, a continuous risk factor and outcome, and under assumptions of linearity and no effect modification, the ratio estimate is defined as: 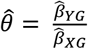, where 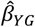 is the coefficient from regressing *Y* on *G*, and 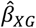 is the coefficient from regressing *X* on *G* [11].

Collider bias will occur when adjusting for a collider variable *C* in the regression models for the ratio estimate, since an association between the instrument and the outcome will occur through conditioning on the collider. To demonstrate the impact and magnitude of collider bias, we performed a simulation study in which we compared estimates when no adjustment on *C* is made versus when the outcome regression is adjusted for *C*. It is also possible to adjust the risk factor regression for *C*; however, while this will distort estimates, this adjustment alone will not bias causal estimates when the true causal effect is null.

### Stratification in Mendelian randomization

To further illustrate the impact of collider bias, we performed a simulation study in which we calculated causal estimates using the ratio method within strata of the population defined using a variable that is influenced by the risk factor, and hence is a collider. We compared two approaches: first, we stratified directly on the collider *C*, and second, we stratified on a new variable *C*_*0*_, referred to as the “residual collider”. The residual collider was generated as the residual from regression of the collider on the genetic instruments:

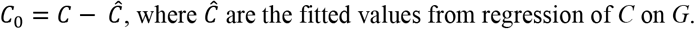

The residual collider *C*_*0*_ is not associated with the instrument, and hence it is not itself a collider. It is influenced by the component of the risk factor that is not a function of *G*, but not by the component that is a function of *G*. However, provided that the genetic instrument does not explain much of the variance in the risk factor (as is typical in a MR application), it is likely not to explain much of the variance in the collider, and so the residual collider will be highly correlated with the collider. Hence, while stratifying on the residual collider is important to avoid bias, the strata defined by stratifying on the collider or residual collider are likely to be similar and so any difference in the interpretation of stratum-specific estimates is minimal.

Here we considered estimates in four strata of the population defined by quartiles of the distribution of the collider or residual collider; however, in practice any number of strata could be considered. We estimated genetic associations with the outcome in each stratum separately. We estimated genetic associations with the risk factor in the full dataset, although if it is believed that these associations vary between strata, it would be possible to estimate these within each stratum as well. The stratum-specific estimate is calculated as the ratio of the stratum-specific genetic association with the outcome divided by the genetic association with the risk factor. We also investigated heterogeneity between the stratum-specific estimates using Cochran’s Q statistic [12], and (in the applied example) we examined the presence of a trend in the estimates by meta-regression of the stratum-specific estimates on the median value of the collider in each stratum [13].

### Simulation set-up

To investigate the impact of collider bias in realistic scenarios, we generated simulated data using the following data-generating model:

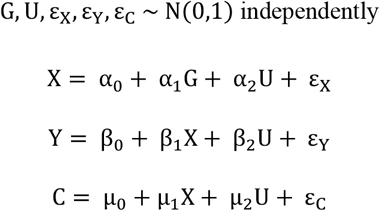

We simulated the instrument *G*, the confounder *U*, and the error terms for *X, Y* and *C*, ε_X_, ε_Y_ and ε_C_, as independent normally distributed variables. The risk factor *X* is defined as a linear combination of the instrument, the confounder, and the error term. The outcome *Y* and the collider *C* are both linear combinations of the risk factor, confounder, and their error terms. In each simulated dataset, we also generated the residual collider *C*_*0*_ as the residual from regression of *C* on *G* as previously described.

The causal estimate of interest is *β*_1_, while *α*_2_ and *β*_2_ represent the effects of *U* on *X* and *Y* respectively; *α*_1_ is the effect of *G* on *X*; and *μ*_1_ and *μ*_2_ are the effects of *X* and *U* on *C*, respectively.

We considered three scenarios based on the parameter *β*_1_: Scenario A1, where there is a null causal effect of *X* on *Y* (β_1_ = 0); Scenario A2, where the effect is constant and positive (β_1_ = 0.5); and Scenario A3, where the effect depends on *C* (β_1_ = 0.5 + 0.2C). In Scenario A1, we considered estimates from the ratio method with and without adjustment for the collider. In Scenarios A2 and A3, we consider stratum-specific estimates from stratification on the collider *C* or the residual collider *C*_*0*_.

We varied the other parameters to consider the impact of different settings on collider bias: i) α_1_ = (0.05, 0.1, and 0.3), in order to study the impact of the strength of the instrument on estimates; ii) positive confounding (α_2_ = 0.8, β_2_ = 0.8) negative (α_2_ = −0.8, β_2_ = −0.8) and mixed (α_2_ = 0.8, β_2_ = −0.8), to study how the direction of confounding affects the estimates and, iii) μ_1_ and μ_2_ = (−1, −0.5, 0, 0.5, 1) to study how the strength of the collider effects influence bias.

We also considered scenarios where the collider is a common effect of *X* and *Y* (Figure 1C). In these scenarios, the collider is generated as C = μ_0_ + μ_1_X + μ_2_U + μ_3_Y + ε_C_, where μ_2_ = 0.3 and μ_3_ = (−1, −0.5, 0, 0.5, 1). In Scenario B1, the causal effect of *X* on *Y* is null (β_1_ = 0), in Scenario B2, the causal effect is constant and positive (β_1_ = 0.5), and in Scenario B3, the causal effect depends on *U* (β_1_ = 0.5 + 0.2*U*), as it is not possible for the causal effect to depend on *C* when *C* is a function of *Y*. Finally, we investigated additional scenarios with a binary outcome *Y*. We generate *Y* from a Binomial distribution where the probability is obtained from a logit transformation as: logit(*P*(Y = 1)) = β_0_ + β_1_X + β_2_U, where β_0_ = 0.5. In Scenario C1, the causal effect of *X* on *Y* is null (β_1_ = 0), in Scenario C2, the causal effect is constant and positive (β_1_ = 0.5) and in Scenario C3, the causal effect depends on *C* (β_1_ = 0.5 + 0.2C). In the binary outcome scenarios, genetic associations with the outcome were estimated by logistic regression. For these additional scenarios, we only consider α_1_ = 0.1 and the positive confounding values; otherwise, we consider all parameters as in scenarios A1 to A3.

We considered a sample size of n = 10,000 and m = 500 replications for each set of parameter values. A directed acyclic graph illustrating the simulation parameters is shown in Figure 1D.

### Applied example: effect of tobacco smoking on bladder cancer risk across bodyweight strata

We applied the proposed MR stratification approach to investigate the causal effect of tobacco smoking on bladder cancer across strata of the population defined by bodyweight. Tobacco smoking is one of the strongest risk factors for cancer, and it has already been reported to be causally associated with bladder cancer risk in a previous Mendelian randomization study [14]. With our current example, the objective was to investigate whether the effect of smoking on the risk of developing bladder cancer is homogeneous across the bodyweight distribution of the population, while avoiding potential collider bias by applying our new stratification approach.

We performed analyses in the UK Biobank study, a population-based cohort of more than 500,000 United Kingdom residents recruited between 2006 and 2010 [15]. For our analysis, we restricted to unrelated European ancestry participants, resulting in a final sample size of 367,643 individuals following sample selection and quality control procedures as described previously [14]. The risk factor is a binary variable representing the smoking behaviour, defined as being a current smoker versus a former or never smoker; the stratifying variable is bodyweight, measured in kg; and the binary outcome is bladder cancer status, defined based on the data from national registries (International Classification of Diseases 9^th^ edition codes: 188, 189.1, 189.2, V10.51, V10.53; or International Classification of Diseases 10^th^ edition codes: C67, C65, C66, Z85.51, Z85.54, Z85.53), and self-reported information from an interview with a nurse practitioner. The instrument for smoking was a weighted genetic risk score comprising 378 conditionally independent SNPs obtained from a genome-wide association study (GWAS) assessing associations with smoking initiation (i.e., probability of ever smoked regularly), and weighted by the associations with smoking initiation [16]. Genetic associations with the risk factor and outcome were obtained by logistic regression in UK Biobank with adjustment for age, sex, and 10 genomic principal components. While age, sex, and principal components cannot logically be colliders as they are not affected by the risk factor or outcome, bodyweight is likely to be a collider, as it is influenced by smoking status [17].

## Results

### Illustration of collider bias

Results from Scenario A1 (β_1_ = 0, null causal effect) are presented in Table 1 for α_1_ = 0.1 (corresponding to R^2^=0.006 for the mean proportion of variance in the risk factor explained by the instrument and a mean F statistic of 60.8) and Supplementary Tables 1 and 2 for α_1_ = 0.3 (corresponding to R^2^=0.051, mean F statistic of 548.6) and α_1_ = 0.05 (corresponding to R^2^=0.001, mean F statistic of 15.3). In each case, we report the median estimate of β_1_ across simulations, and the empirical type I error rate, representing the proportion of simulated datasets where the 95% confidence interval for the ratio estimate excludes zero. With no adjustment for the collider, median estimates were close to zero and empirical type I error rate was close to the expected value of 5%. When adjusting for the collider in the regression of *Y* on *G*, estimates were biased, and type I error rates were substantially above 5%. The only exception was for μ_1_ = 0; in this case, the variable *C* is not a function of the risk factor, and so does not act as a collider. Bias and type I error rates generally increased for more extreme values of μ_1_ and μ_2_ (both positive and negative values). The direction of bias depended on μ_1_ and μ_2_ and the direction of confounding.

**Table 1.** Median of β_1_ estimates and empirical Type I error rates for Scenario A1 (null causal effect, β_1_ =0) with positive, negative, and mixed confounding, and α_1_ =0.1

| | | Positive confounding ($\alpha_2$ and $\beta_2 = 0.8$) | | | | Negative confounding ($\alpha_2$ and $\beta_2 = -0.8$) | | | | Mixed confounding ($\alpha_2 = 0.8$ and $\beta_2 = -0.8$) | | | |
| --- | --- | --- | --- | --- | --- | --- | --- | --- | --- | --- | --- | --- | --- |
|  |  | Median estimate | Type I error rate (%) | Median estimate | Type I error rate (%) | Median estimate | Type I error rate (%) | Median estimate | Type I error rate (%) | Median estimate | Type I error rate (%) | Median estimate | Type I error rate (%) |
| $\mu_1$ | $\mu_2$ | No adjust for collider | | Adjust Y/G for collider | | No adjust for collider | | Adjust Y/G for collider | | No adjust for collider | | Adjust Y/G for collider | |
| -1 | -1 | 0.01 | 7% | -0.27 | 70% | 0.01 | 5% | 0.09 | 10% | 0.00 | 5% | 0.28 | 69% |
|  | -0.5 | 0.00 | 6% | -0.28 | 66% | -0.01 | 3% | -0.12 | 15% | 0.00 | 6% | 0.29 | 69% |
|  | 0 | 0.00 | 5% | -0.24 | 50% | -0.01 | 4% | -0.26 | 53% | 0.01 | 5% | 0.25 | 54% |
|  | 0.5 | 0.01 | 6% | -0.10 | 15% | 0.01 | 6% | -0.28 | 69% | 0.00 | 6% | 0.12 | 15% |
|  | 1 | 0.00 | 5% | 0.08 | 8% | 0.01 | 3% | -0.27 | 68% | 0.00 | 4% | -0.08 | 11% |
| -0.5 | -1 | 0.01 | 6% | -0.16 | 30% | 0.00 | 6% | 0.15 | 21% | 0.00 | 5% | 0.17 | 34% |
|  | -0.5 | 0.00 | 6% | -0.18 | 32% | 0.00 | 5% | 0.04 | 7% | 0.01 | 4% | 0.18 | 30% |
|  | 0 | 0.00 | 5% | -0.11 | 16% | 0.00 | 3% | -0.12 | 14% | 0.00 | 3% | 0.11 | 14% |
|  | 0.5 | -0.01 | 5% | 0.02 | 5% | 0.00 | 6% | -0.17 | 30% | 0.00 | 4% | -0.03 | 7% |
|  | 1 | 0.01 | 5% | 0.15 | 25% | 0.00 | 6% | -0.18 | 34% | 0.01 | 4% | -0.15 | 21% |
| 0 | -1 | -0.01 | 7% | 0.00 | 6% | 0.00 | 6% | 0.00 | 6% | 0.00 | 6% | 0.00 | 6% |
|  | -0.5 | 0.00 | 7% | 0.00 | 6% | 0.00 | 6% | 0.00 | 6% | 0.00 | 5% | -0.01 | 5% |
|  | 0 | 0.00 | 6% | 0.01 | 6% | 0.01 | 6% | 0.01 | 6% | 0.00 | 6% | 0.00 | 6% |
|  | 0.5 | -0.01 | 6% | 0.00 | 6% | 0.00 | 5% | 0.00 | 6% | 0.00 | 7% | 0.00 | 7% |
|  | 1 | 0.00 | 5% | 0.01 | 6% | 0.00 | 4% | 0.00 | 4% | -0.01 | 6% | -0.01 | 5% |
| 0.5 | -1 | 0.01 | 4% | 0.16 | 22% | 0.01 | 6% | -0.17 | 35% | 0.00 | 4% | -0.15 | 23% |
|  | -0.5 | -0.01 | 5% | 0.02 | 5% | -0.01 | 7% | -0.19 | 36% | 0.01 | 6% | -0.03 | 7% |
|  | 0 | 0.00 | 5% | -0.11 | 14% | 0.00 | 4% | -0.11 | 15% | -0.01 | 4% | 0.10 | 15% |
|  | 0.5 | 0.01 | 4% | -0.17 | 28% | 0.00 | 5% | 0.03 | 7% | 0.00 | 5% | 0.18 | 34% |
|  | 1 | 0.01 | 6% | -0.17 | 31% | 0.01 | 5% | 0.15 | 23% | 0.00 | 5% | 0.17 | 33% |
| 1 | -1 | 0.01 | 5% | 0.08 | 8% | 0.01 | 5% | -0.27 | 70% | 0.01 | 3% | -0.07 | 9% |
|  | -0.5 | 0.01 | 4% | -0.10 | 13% | 0.01 | 5% | -0.27 | 64% | -0.01 | 5% | 0.11 | 18% |
|  | 0 | 0.01 | 6% | -0.24 | 52% | 0.00 | 5% | -0.24 | 50% | -0.01 | 3% | 0.24 | 48% |
|  | 0.5 | 0.01 | 5% | -0.27 | 66% | 0.01 | 4% | -0.11 | 15% | 0.00 | 4% | 0.28 | 66% |
|  | 1 | 0.01 | 4% | -0.26 | 68% | 0.00 | 4% | 0.08 | 10% | 0.02 | 5% | 0.29 | 75% |

- Empirical Type I error rate represents the proportion of simulated datasets where the null hypothesis is not rejected.

### Stratification in Mendelian randomization

Results from Scenario A2 (β_1_ = 0.5, constant positive effect) are presented in Table 2 for α_1_ = 0.1 with positive confounding. Supplementary Table 3 shows results for α_1_ = 0.1 with negative and mixed confounding, and Supplementary Tables 4 and 5 for α_1_ = 0.3 and α_1_ = 0.05. We report the median estimate of β_1_ in four strata of the sample defined by quartiles of the collider *C* or residual collider *C*_*0*_, and the proportion of simulated datasets for which the heterogeneity test statistic is rejected. When stratifying on the collider, median estimates were somewhat variable between the strata, although the proportion of datasets in which the heterogeneity test rejects the null hypothesis of homogeneity was not much above 5% in any scenario, reaching a maximum of 11% when α_1_ = 0.3. However, if we considered stronger instruments or larger sample sizes, we would see this proportion considerably exceed 5% (see Supplementary Table 6 where we first set α_1_ = 0.5 and n=10,000, and then set α_1_ = 0.1 and n = 50,000, and the type I error rate reached 16% in each case). Median estimates differed substantially from the true value of 0.5 across strata, especially when the collider was strongly affected by the risk factor. In contrast, when stratifying on the residual collider, median estimates of β_1_ were close to 0.5 throughout, and there was no suggestion in any case that the heterogeneity test rejected the null above the expected 5% rate.

**Table 2.** Median of causal estimates in different quartiles, and proportion of datasets in which the homogeneity test was rejected for Scenario A2 (fixed causal effect of β_1_ =0.5) with positive confounding and α_1_ =0.1

| | | Positive confounding ($\alpha_2$ and $\beta_2 = 0.8$) | | | | | | | | | |
| --- | --- | --- | --- | --- | --- | --- | --- | --- | --- | --- | --- |
|  |  | Stratifying on collider, C |  |  |  |  | Stratifying on residual collider, C<sub>0</sub> |  |  |  |  |
| $\mu_1$ | $\mu_2$ | Proportion homogeneity rejected (%) | Median estimates Q1 | Median estimates Q2 | Median estimates Q3 | Median estimates Q4 | Proportion homogeneity rejected (%) | Median estimates Q1 | Median estimates Q2 | Median estimates Q3 | Median estimates Q4 |
| -1 | -1 | 8% | 0.11 | -0.01 | 0.01 | 0.10 | 8% | 0.49 | 0.48 | 0.49 | 0.50 |
|  | -0.5 | 6% | 0.09 | -0.05 | -0.05 | 0.09 | 5% | 0.53 | 0.48 | 0.51 | 0.53 |
|  | 0 | 7% | 0.07 | 0.01 | -0.04 | 0.07 | 6% | 0.50 | 0.52 | 0.50 | 0.49 |
|  | 0.5 | 7% | 0.19 | 0.10 | 0.09 | 0.17 | 6% | 0.49 | 0.50 | 0.49 | 0.48 |
|  | 1 | 4% | 0.41 | 0.37 | 0.40 | 0.40 | 4% | 0.50 | 0.48 | 0.52 | 0.49 |
| -0.5 | -1 | 7% | 0.30 | 0.21 | 0.23 | 0.26 | 5% | 0.53 | 0.53 | 0.52 | 0.48 |
|  | -0.5 | 5% | 0.24 | 0.16 | 0.18 | 0.25 | 5% | 0.48 | 0.47 | 0.51 | 0.48 |
|  | 0 | 4% | 0.29 | 0.23 | 0.21 | 0.29 | 4% | 0.50 | 0.47 | 0.45 | 0.49 |
|  | 0.5 | 6% | 0.47 | 0.42 | 0.47 | 0.46 | 6% | 0.50 | 0.50 | 0.50 | 0.50 |
|  | 1 | 3% | 0.59 | 0.65 | 0.64 | 0.60 | 5% | 0.48 | 0.50 | 0.51 | 0.50 |
| 0 | -1 | 6% | 0.50 | 0.50 | 0.48 | 0.48 | 6% | 0.51 | 0.50 | 0.48 | 0.47 |
|  | -0.5 | 4% | 0.48 | 0.48 | 0.52 | 0.50 | 4% | 0.47 | 0.47 | 0.52 | 0.50 |
|  | 0 | 4% | 0.49 | 0.48 | 0.51 | 0.52 | 4% | 0.49 | 0.49 | 0.51 | 0.51 |
|  | 0.5 | 5% | 0.52 | 0.49 | 0.49 | 0.53 | 6% | 0.54 | 0.49 | 0.48 | 0.53 |
|  | 1 | 4% | 0.48 | 0.48 | 0.51 | 0.49 | 4% | 0.48 | 0.47 | 0.52 | 0.50 |
| 0.5 | -1 | 4% | 0.62 | 0.63 | 0.66 | 0.63 | 4% | 0.51 | 0.49 | 0.52 | 0.52 |
|  | -0.5 | 5% | 0.45 | 0.44 | 0.44 | 0.47 | 4% | 0.49 | 0.50 | 0.49 | 0.51 |
|  | 0 | 4% | 0.32 | 0.23 | 0.23 | 0.29 | 4% | 0.51 | 0.51 | 0.47 | 0.49 |
|  | 0.5 | 5% | 0.25 | 0.18 | 0.19 | 0.25 | 3% | 0.51 | 0.49 | 0.52 | 0.50 |
|  | 1 | 5% | 0.26 | 0.23 | 0.20 | 0.28 | 5% | 0.46 | 0.51 | 0.49 | 0.50 |
| 1 | -1 | 5% | 0.41 | 0.37 | 0.34 | 0.40 | 5% | 0.49 | 0.50 | 0.46 | 0.49 |
|  | -0.5 | 5% | 0.16 | 0.11 | 0.09 | 0.21 | 4% | 0.49 | 0.49 | 0.50 | 0.51 |
|  | 0 | 6% | 0.12 | -0.03 | 0.00 | 0.11 | 6% | 0.50 | 0.50 | 0.53 | 0.51 |
|  | 0.5 | 6% | 0.04 | -0.03 | -0.02 | 0.07 | 4% | 0.47 | 0.51 | 0.51 | 0.50 |
|  | 1 | 6% | 0.14 | 0.00 | 0.03 | 0.10 | 6% | 0.54 | 0.49 | 0.50 | 0.49 |

Proportion homogeneity rejected represents the proportion of simulated datasets where the null hypothesis of homogeneity is rejected

Results from Scenario A3 (variable effect) are presented in Table 3 for α_1_ = 0.1 with positive confounding. Supplementary Table 7 shows results for α_1_ = 0.1 with negative and mixed confounding, and Supplementary Tables 8 and 9 for α_1_ = 0.3 and α_1_ = 0.05. Estimates differed somewhat when stratifying on the collider versus the residual collider, although in both cases median estimates increased across the four strata. The proportion of datasets in which the heterogeneity test was rejected, which in this case represents the empirical power to detect heterogeneity in the stratum-specific estimates, was consistently higher when stratifying on the residual collider, indicating that true differences in the stratum-specific estimates were better detected when stratifying on the residual collider.

**Table 3.** Median of causal estimates in different quartiles, and proportion of datasets in which the homogeneity test was rejected for Scenario A3 (varying causal effect) with positive confounding and α_1_ =0.1

| | | Positive confounding ($\alpha_2$ and $\beta_2 = 0.8$) | | | | | | | | | |
| --- | --- | --- | --- | --- | --- | --- | --- | --- | --- | --- | --- |
|  |  | Stratifying on collider, C |  |  |  |  | Stratifying on residual collider, C<sub>0</sub> |  |  |  |  |
| $\mu_1$ | $\mu_2$ | Proportion<br>homogeneity<br>rejected (%) | Median<br>estimates<br>Q1 | Median<br>estimates<br>Q2 | Median<br>estimates<br>Q3 | Median<br>estimates<br>Q4 | Proportion<br>homogeneity<br>rejected (%) | Median<br>estimates<br>Q1 | Median<br>estimates<br>Q2 | Median<br>estimates<br>Q3 | Median<br>estimates<br>Q4 |
| -1 | -1 | 48% | -0.39 | -0.07 | 0.14 | 0.58 | 95% | -0.46 | 0.19 | 0.64 | 1.27 |
|  | -0.5 | 30% | -0.33 | -0.06 | 0.00 | 0.44 | 88% | -0.36 | 0.24 | 0.59 | 1.15 |
|  | 0 | 19% | -0.25 | -0.08 | 0.00 | 0.38 | 78% | -0.25 | 0.22 | 0.55 | 1.09 |
|  | 0.5 | 15% | -0.11 | 0.05 | 0.14 | 0.46 | 61% | -0.19 | 0.24 | 0.56 | 0.99 |
|  | 1 | 16% | 0.09 | 0.32 | 0.44 | 0.63 | 40% | -0.09 | 0.28 | 0.54 | 0.88 |
| -0.5 | -1 | 36% | -0.11 | 0.15 | 0.32 | 0.71 | 68% | -0.07 | 0.34 | 0.64 | 1.08 |
|  | -0.5 | 19% | -0.07 | 0.14 | 0.30 | 0.58 | 48% | -0.01 | 0.38 | 0.64 | 0.94 |
|  | 0 | 14% | 0.08 | 0.23 | 0.36 | 0.57 | 25% | 0.11 | 0.41 | 0.59 | 0.87 |
|  | 0.5 | 11% | 0.22 | 0.46 | 0.55 | 0.76 | 16% | 0.16 | 0.43 | 0.56 | 0.83 |
|  | 1 | 16% | 0.35 | 0.58 | 0.77 | 0.98 | 16% | 0.19 | 0.40 | 0.57 | 0.81 |
| 0 | -1 | 24% | 0.24 | 0.49 | 0.66 | 0.95 | 24% | 0.24 | 0.51 | 0.70 | 0.94 |
|  | -0.5 | 14% | 0.33 | 0.52 | 0.67 | 0.90 | 15% | 0.33 | 0.50 | 0.66 | 0.89 |
|  | 0 | 13% | 0.34 | 0.52 | 0.66 | 0.86 | 13% | 0.35 | 0.53 | 0.66 | 0.87 |
|  | 0.5 | 13% | 0.33 | 0.51 | 0.69 | 0.88 | 13% | 0.33 | 0.52 | 0.69 | 0.89 |
|  | 1 | 25% | 0.24 | 0.51 | 0.69 | 0.98 | 26% | 0.25 | 0.52 | 0.70 | 0.99 |
| 0.5 | -1 | 18% | 0.45 | 0.69 | 0.87 | 1.07 | 18% | 0.38 | 0.59 | 0.77 | 1.01 |
|  | -0.5 | 15% | 0.34 | 0.50 | 0.64 | 0.88 | 18% | 0.37 | 0.61 | 0.79 | 1.03 |
|  | 0 | 14% | 0.17 | 0.30 | 0.41 | 0.68 | 26% | 0.30 | 0.61 | 0.77 | 1.07 |
|  | 0.5 | 19% | 0.05 | 0.22 | 0.37 | 0.72 | 45% | 0.20 | 0.57 | 0.84 | 1.17 |
|  | 1 | 34% | 0.00 | 0.25 | 0.41 | 0.81 | 63% | 0.13 | 0.56 | 0.83 | 1.27 |
| 1 | -1 | 16% | 0.25 | 0.46 | 0.55 | 0.88 | 40% | 0.26 | 0.68 | 0.90 | 1.33 |
|  | -0.5 | 12% | 0.03 | 0.16 | 0.30 | 0.58 | 53% | 0.19 | 0.65 | 0.97 | 1.37 |
|  | 0 | 18% | -0.10 | -0.02 | 0.11 | 0.49 | 70% | 0.13 | 0.63 | 0.97 | 1.46 |
|  | 0.5 | 23% | -0.16 | -0.02 | 0.11 | 0.60 | 83% | 0.04 | 0.59 | 0.98 | 1.54 |
|  | 1 | 35% | -0.23 | 0.01 | 0.20 | 0.71 | 94% | -0.07 | 0.55 | 1.04 | 1.65 |

3 Proportion homogeneity rejected represents the proportion of simulated datasets where the null hypothesis of homogeneity is rejected

### Additional scenarios

In Scenarios B1 (β_1_ = 0), B2 (β_1_ = 0.5) and B3 (β_1_ = 0.5 + 0.2U), where the collider was a function of both the risk factor and outcome, similar results were observed, with collider bias evident when conditioning on the collider (Supplementary Table 10) and when stratifying on the collider (Supplementary Table 11). Collider bias in Scenarios B1 and B2 was greater compared with Scenarios A1 and A2 where the collider was a function of the risk factor only. Similarly, bias was not observed when stratifying on the residual collider (Supplementary Table 11). For Scenario B3, the power of the homogeneity test was lower in comparison to Scenario A3 (Supplementary Table 11), as the dependence of effect heterogeneity on the collider was weaker; however, heterogeneity was detected more often when stratifying on the residual collider than on the collider.

For Scenarios C1 (β_1_ = 0), C2 (β_1_ = 0.5) and C3 (β_1_ = 0.5 + 0.2C), where the outcome was binary, again similar results were observed, with collider bias evident when conditioning on the collider in Scenario C1 (Supplementary Table 12) and when stratifying on the collider in Scenarios C2 and C3 (Supplementary Table 13). Bias was smaller than in cases with a continuous outcome, although direct comparison is somewhat unfair as estimates with a binary outcome were obtained from logistic regression and so represent log odds ratios. Estimates when stratifying on the residual collider were slightly attenuated from 0.5 due to the non-collapsibility of the odds ratio [18,19]. Despite this, in Scenario C2 we observed similar estimates across the different strata of *C*_*0*_ for each set of parameter values. Similarly, in Scenario C3 we observed that median stratum-specific estimates increased across the four strata when stratifying on either the collider or residual collider. Power to detect heterogeneity was lower compared with Scenario A3 as the stratum-specific estimates are less precise, although again power was consistently higher when stratifying on the residual collider.

### Applied example: effect of tobacco smoking on bladder cancer risk across bodyweight strata

Estimates for the causal effect of smoking on bladder cancer in strata of bodyweight and residual bodyweight are shown in Table 4. Estimates represent the odds ratio for bladder cancer per one unit increase in the log odds of being a current smoker. Estimates were positive in all strata, although larger in strata 1 and 2 for both bodyweight and residual bodyweight, and 95% confidence intervals excluded the null in these strata only. Although the homogeneity test was not rejected for either collider variable (p-value = 0.151 and p-value = 0.084 for bodyweight and residual bodyweight, respectively), there was evidence of trend in the stratum-specific estimates for residual bodyweight from meta-regression on the mean value of bodyweight in each stratum (p-value = 0.019). These results suggest that the effect of smoking on bladder cancer is stronger for subgroups of the population with lower bodyweight.

**Table 4.**
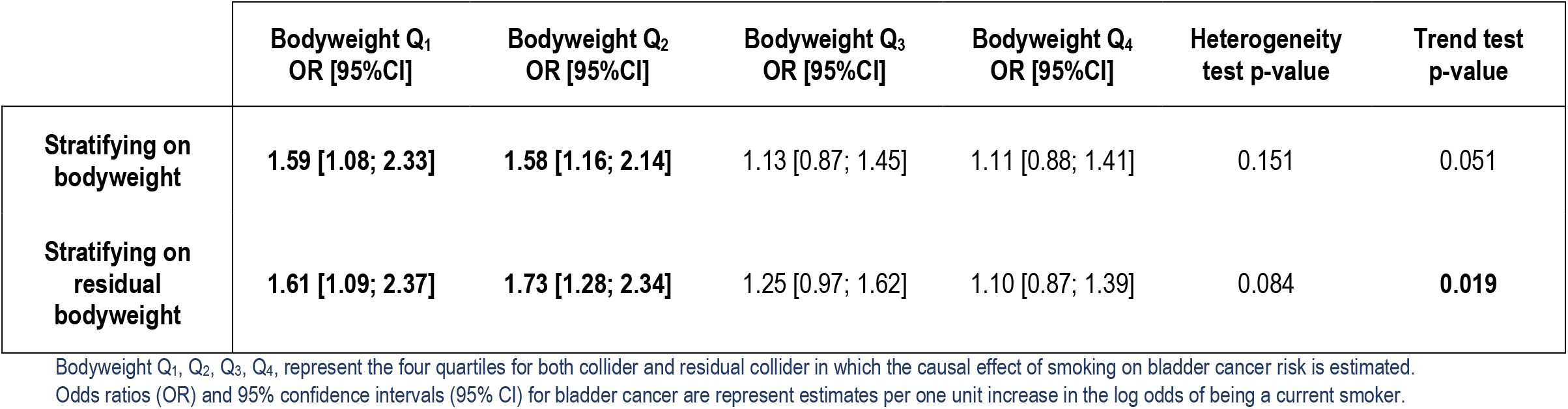
Applied example using UK Biobank to investigate the effect of smoking status on bladder cancer risk in different bodyweight strata.

## Discussion

In this paper, we have demonstrated that conditioning or stratifying on a variable that is a collider can have a serious impact on MR estimates. We have introduced a simple approach that constructs a new variable, the residual collider, which is typically highly correlated with the collider, but is independent of the instrument. Estimates obtained from stratification on the residual collider did not suffer from bias in a range of simulation studies. Stratification on the residual collider allows investigators to explore causal estimation in relevant subgroups of the population. We applied our new approach to demonstrate that MR estimates for the effect of smoking on bladder cancer differ within strata of bodyweight, suggesting that the effect of smoking is stronger for subgroups of the population with lower bodyweight.

The approach of stratifying on the residual collider follows the same logic as a previously proposed method for non-linear MR, in which causal estimates are obtained in strata of the population defined by the “residual risk factor” or “IV-free exposure” [20,21]. This variable is defined similarly to the residual collider, except the collider variable is the risk factor itself. This method has been used previously to estimate the causal effect of blood pressure on coronary heart disease risk within strata of blood pressure, resulting in a curve that represents the shape of the causal relationship between the risk factor and the outcome [22]. This paper extends on that method, showing that the same idea can be used to provide causal estimates stratified on a separate variable even if that variable is a collider.

There are some limitations to this approach. First, while the independence of the residual collider from the instrument is theoretically justified, we demonstrated the validity of our approach through simulation studies. Although we considered a range of different scenarios and parameter values, it is not possible to consider every possible data-generating mechanism by which that a collider could arise. Second, in practice, the relationships between variables are unknown, and so it may be unclear whether a proposed stratifying variable is a collider. However, even if the variable is not a collider, it is unlikely stratification on the residual variable will lead to invalid estimates, suggesting that this approach would be valid for stratifying on variables that are not colliders. This was demonstrated in the simulation study when the effect of the risk factor on the “collider” was zero (μ_1_ = 0), and so the stratifying variable was not a collider. One exception is if the stratifying variable is on the causal pathway from the risk factor to the outcome. Stratification on such a variable (a “mediator”) will lead to biased estimates even in the proposed approach. Finally, the degree of collider bias depended on the strength of the effects of the risk factor and confounder on the collider, and the direction of confounding. It is possible that collider bias may not be substantial in practice, as observed in the applied example, where estimates were broadly similar when stratifying on bodyweight or residual bodyweight. However, the power to detect heterogeneity in stratum-specific estimates in the simulation study was greater when stratifying on the residual collider, especially when the proportion of variance of the risk factor explained by the instrument was higher. This was also observed in the applied example, where a lower p-value was observed in both the heterogeneity test and the trend test when stratifying on residual bodyweight.

The finding that the effect of smoking on bladder cancer is greater in lower bodyweight subgroups is plausible, because for any given level of cigarette consumption smaller individuals will tend to be exposed to greater concentrations of carcinogens [23]. An alternative explanation is that the genetic variants could associate more strongly with smoking intensity in individuals of lower bodyweight. However, we would be cautious not to interpret estimates in the higher bodyweight quartiles as implying an absence of a causal effect in heavier individuals; it is possible that the null estimates reflect limited power. A limitation of the applied example is overlap between the discovery dataset for the genetic variants, and the dataset used in the MR analysis, which can lead to winner’s curse, and the one-sample setting, which can lead to weak instrument bias.

In conclusion, we recommend that researchers performing MR to investigate causal effects in strata of a population defined by a collider stratify on residual values of the collider rather than stratifying on the collider directly.

## Data Availability

Not available

## Funding

SB is supported by a Sir Henry Dale Fellowship jointly funded by the Wellcome Trust and the Royal Society (204623/Z/16/Z). This research was funded by United Kingdom Research and Innovation Medical Research Council (MC_UU_00002/7) and supported by the National Institute for Health Research Cambridge Biomedical Research Centre (BRC-1215-20014). The views expressed are those of the authors and not necessarily those of the National Institute for Health Research or the Department of Health and Social Care.

The work was partially supported by Fondo de Investigaciones Sanitarias (FIS), Instituto de Salud Carlos III, Spain (#PI18/01347), Ministerio de Ciencia e Innovación, Spain (#PID2019-104681RB-I00). CC received a fellowship from CIBERONC, Insitituto de Salud Carlos III, Spain.

## Competing interests

DG is employed part time by Novo Nordisk, outside of the submitted work

