## Supplementary material for "Avoiding bias in Mendelian randomization when stratifying on a collider"

### Supplementary Tables

**Supplementary Table 1.** Scenario A1, results for positive, negative, and mixed confounding, adjustment approach, $\alpha_1$=0.3

| | | Positive confounding ($\alpha_2$ and $\beta_2$ = 0.8) | | Negative confounding ($\alpha_2$ and $\beta_2$ = -0.8) | | Mixed confounding ($\alpha_2$ =0.8 and $\beta_2$ = -0.8) | |
|---|---|---|---|---|---|---|---|
| | | Median of causal estimates | Type I error rate (%) | Median of causal estimates | Type I error rate (%) | Median of causal estimates | Type I error rate (%) |
| $\mu_1$ | $\mu_2$ | *No adjust for collider* | | | | | |
| -0.5 | -0.5 | 0.00 | 6% | 0.00 | 5% | 0.00 | 6% |
| | 0 | 0.01 | 4% | 0.00 | 6% | 0.00 | 2% |
| | 0.5 | 0.00 | 5% | 0.00 | 4% | 0.00 | 5% |
| 0 | -0.5 | 0.00 | 4% | 0.00 | 4% | 0.00 | 4% |
| | 0 | 0.00 | 5% | 0.00 | 5% | 0.00 | 6% |
| | 0.5 | 0.00 | 4% | 0.00 | 5% | 0.00 | 5% |
| 0.5 | -0.5 | 0.00 | 4% | 0.00 | 5% | 0.00 | 5% |
| | 0 | 0.00 | 7% | 0.00 | 4% | 0.00 | 4% |
| | 0.5 | 0.00 | 6% | 0.00 | 6% | 0.00 | 4% |
| $\mu_1$ | $\mu_2$ | *Adjust Y/G for collider* | | | | | |
| -0.5 | -0.5 | -0.17 | 100% | 0.04 | 13% | 0.18 | 100% |
| | 0 | -0.12 | 81% | -0.12 | 77% | 0.11 | 76% |
| | 0.5 | 0.03 | 11% | -0.18 | 100% | -0.03 | 10% |
| 0 | -0.5 | 0.00 | 6% | 0.00 | 5% | 0.00 | 6% |
| | 0 | 0.00 | 4% | 0.00 | 5% | 0.00 | 5% |
| | 0.5 | 0.00 | 4% | 0.00 | 4% | 0.00 | 6% |
| 0.5 | -0.5 | 0.04 | 13% | -0.18 | 99% | -0.03 | 12% |
| | 0 | -0.11 | 75% | -0.11 | 76% | 0.11 | 77% |
| | 0.5 | -0.17 | 99% | 0.03 | 9% | 0.18 | 100% |

- Empirical Type I error rate represents the proportion of simulated datasets where the null hypothesis is not rejected

**Supplementary Table 2.** Scenario A1, results for positive, negative, and mixed confounding, adjustment approach, $\alpha_1$=0.05

| | | Positive confounding ($\alpha_2$ and $\beta_2$ = 0.8) | | Negative confounding ($\alpha_2$ and $\beta_2$ = -0.8) | | Mixed confounding ($\alpha_2$ =0.8 and $\beta_2$ = -0.8) | |
|---|---|---|---|---|---|---|---|
| | | Median of causal estimates | Type I error rate (%) | Median of causal estimates | Type I error rate (%) | Median of causal estimates | Type I error rate (%) |
| $\mu_1$ | $\mu_2$ | *No adjust for collider* | | | | | |
| -0.5 | -0.5 | -0.02 | 3% | 0.00 | 7% | -0.03 | 5% |
| | 0 | -0.01 | 5% | 0.01 | 5% | -0.01 | 4% |
| | 0.5 | -0.01 | 5% | 0.00 | 5% | 0.03 | 5% |
| 0 | -0.5 | 0.05 | 5% | 0.00 | 3% | 0.00 | 6% |
| | 0 | -0.01 | 4% | -0.01 | 5% | -0.02 | 5% |
| | 0.5 | 0.02 | 5% | 0.00 | 5% | -0.04 | 6% |
| 0.5 | -0.5 | 0.00 | 7% | 0.00 | 5% | 0.00 | 5% |
| | 0 | -0.01 | 4% | -0.01 | 5% | -0.01 | 4% |
| | 0.5 | -0.01 | 5% | 0.03 | 3% | 0.00 | 4% |
| $\mu_1$ | $\mu_2$ | *Adjust Y/G for collider* | | | | | |
| -0.5 | -0.5 | -0.18 | 11% | 0.04 | 5% | 0.16 | 13% |
| | 0 | -0.13 | 8% | -0.11 | 6% | 0.13 | 7% |
| | 0.5 | 0.02 | 5% | -0.18 | 13% | -0.05 | 6% |
| 0 | -0.5 | -0.01 | 4% | -0.02 | 6% | -0.02 | 5% |
| | 0 | 0.02 | 5% | 0.00 | 5% | -0.04 | 6% |
| | 0.5 | 0.00 | 6% | 0.01 | 7% | -0.01 | 5% |
| 0.5 | -0.5 | 0.02 | 5% | -0.19 | 13% | -0.04 | 4% |
| | 0 | -0.12 | 7% | -0.08 | 6% | 0.11 | 8% |
| | 0.5 | -0.16 | 9% | 0.03 | 5% | 0.17 | 12% |

Empirical Type I error rate represents the proportion of simulated datasets where the null hypothesis is not rejected

**Supplementary Table 3.** Scenario A2, results for negative, and mixed confounding, stratification approach, $\alpha_1$=0.1

| | | **Negative confounding ($\alpha_2$ and $\beta_2$ = -0.8)** | | | | | | | | | |
|---|---|---|---|---|---|---|---|---|---|---|---|
| | | **Stratifying on collider, C** | | | | | **Stratifying on residual collider, $C_0$** | | | | |
| **$\mu_1$** | **$\mu_2$** | **Proportion homogeneity rejected (%)** | **Median estimates Q1** | **Median estimates Q2** | **Median estimates Q3** | **Median estimates Q4** | **Proportion homogeneity rejected (%)** | **Median estimates Q1** | **Median estimates Q2** | **Median estimates Q3** | **Median estimates Q4** |
| -0.5 | -0.5 | 6% | 0.44 | 0.45 | 0.46 | 0.45 | 3% | 0.48 | 0.48 | 0.51 | 0.52 |
| | 0 | 4% | 0.28 | 0.25 | 0.24 | 0.32 | 5% | 0.49 | 0.49 | 0.49 | 0.5 |
| | 0.5 | 6% | 0.26 | 0.17 | 0.19 | 0.22 | 6% | 0.52 | 0.49 | 0.5 | 0.46 |
| 0 | -0.5 | 4% | 0.51 | 0.51 | 0.50 | 0.49 | 4% | 0.52 | 0.51 | 0.49 | 0.49 |
| | 0 | 7% | 0.49 | 0.50 | 0.50 | 0.49 | 5% | 0.5 | 0.49 | 0.5 | 0.49 |
| | 0.5 | 4% | 0.47 | 0.50 | 0.49 | 0.48 | 5% | 0.47 | 0.49 | 0.5 | 0.48 |
| 0.5 | -0.5 | 7% | 0.23 | 0.18 | 0.18 | 0.24 | 7% | 0.47 | 0.48 | 0.51 | 0.49 |
| | 0 | 4% | 0.28 | 0.27 | 0.25 | 0.32 | 5% | 0.47 | 0.5 | 0.5 | 0.51 |
| | 0.5 | 6% | 0.46 | 0.44 | 0.46 | 0.47 | 6% | 0.48 | 0.5 | 0.49 | 0.53 |
| | | **Mixed confounding ($\alpha_2$ =0.8 and $\beta_2$ = -0.8)** | | | | | | | | | |
| -0.5 | -0.5 | 6% | 0.54 | 0.53 | 0.52 | 0.52 | 5% | 0.52 | 0.51 | 0.49 | 0.49 |
| | 0 | 6% | 0.47 | 0.45 | 0.47 | 0.47 | 5% | 0.51 | 0.48 | 0.48 | 0.50 |
| | 0.5 | 5% | 0.43 | 0.39 | 0.38 | 0.39 | 5% | 0.51 | 0.51 | 0.48 | 0.49 |
| 0 | -0.5 | 7% | 0.47 | 0.50 | 0.50 | 0.50 | 7% | 0.47 | 0.50 | 0.50 | 0.49 |
| | 0 | 5% | 0.50 | 0.49 | 0.47 | 0.49 | 4% | 0.49 | 0.49 | 0.48 | 0.48 |
| | 0.5 | 5% | 0.50 | 0.52 | 0.51 | 0.50 | 5% | 0.50 | 0.51 | 0.51 | 0.49 |
| 0.5 | -0.5 | 5% | 0.42 | 0.39 | 0.41 | 0.43 | 6% | 0.51 | 0.51 | 0.50 | 0.54 |
| | 0 | 5% | 0.46 | 0.46 | 0.48 | 0.47 | 5% | 0.49 | 0.49 | 0.51 | 0.50 |
| | 0.5 | 5% | 0.51 | 0.52 | 0.54 | 0.54 | 4% | 0.48 | 0.50 | 0.51 | 0.51 |

Proportion homogeneity rejected represents the proportion of simulated datasets where the null hypothesis of homogeneity is rejected

**Supplementary Table 4.** Scenario A2, results for positive, negative, and mixed confounding, stratification approach, $\alpha_1$=0.3

| | | Positive confounding ($\alpha_2$ and $\beta_2$ = 0.8) | | | | | | | | | |
|---|---|---|---|---|---|---|---|---|---|---|---|
| | | Stratifying on collider, C | | | | | Stratifying on residual collider, $C_0$ | | | | |
| $\mu_1$ | $\mu_2$ | Proportion homogeneity rejected (%) | Median estimates Q1 | Median estimates Q2 | Median estimates Q3 | Median estimates Q4 | Proportion homogeneity rejected (%) | Median estimates Q1 | Median estimates Q2 | Median estimates Q3 | Median estimates Q4 |
| -0.5 | -0.5 | 11% | 0.25 | 0.19 | 0.19 | 0.26 | 5% | 0.50 | 0.49 | 0.50 | 0.50 |
| | 0 | 7% | 0.31 | 0.25 | 0.25 | 0.30 | 7% | 0.51 | 0.50 | 0.49 | 0.50 |
| | 0.5 | 5% | 0.47 | 0.44 | 0.46 | 0.46 | 5% | 0.50 | 0.50 | 0.50 | 0.50 |
| 0 | -0.5 | 4% | 0.51 | 0.50 | 0.50 | 0.50 | 5% | 0.51 | 0.50 | 0.50 | 0.50 |
| | 0 | 5% | 0.50 | 0.51 | 0.50 | 0.50 | 5% | 0.50 | 0.51 | 0.50 | 0.50 |
| | 0.5 | 6% | 0.51 | 0.49 | 0.50 | 0.50 | 6% | 0.50 | 0.49 | 0.50 | 0.50 |
| 0.5 | -0.5 | 6% | 0.47 | 0.46 | 0.45 | 0.46 | 5% | 0.51 | 0.51 | 0.50 | 0.50 |
| | 0 | 8% | 0.30 | 0.25 | 0.25 | 0.30 | 6% | 0.50 | 0.51 | 0.50 | 0.49 |
| | 0.5 | 6% | 0.26 | 0.20 | 0.18 | 0.25 | 4% | 0.50 | 0.50 | 0.49 | 0.50 |
| | | Negative confounding ($\alpha_2$ and $\beta_2$ = -0.8) | | | | | | | | | |
| -0.5 | -0.5 | 6% | 0.46 | 0.45 | 0.46 | 0.46 | 5% | 0.49 | 0.50 | 0.50 | 0.50 |
| | 0 | 7% | 0.31 | 0.24 | 0.25 | 0.31 | 5% | 0.50 | 0.50 | 0.50 | 0.50 |
| | 0.5 | 8% | 0.25 | 0.19 | 0.18 | 0.25 | 6% | 0.49 | 0.50 | 0.50 | 0.50 |
| 0 | -0.5 | 3% | 0.50 | 0.51 | 0.49 | 0.49 | 4% | 0.50 | 0.50 | 0.49 | 0.50 |
| | 0 | 4% | 0.50 | 0.51 | 0.49 | 0.50 | 4% | 0.50 | 0.50 | 0.49 | 0.50 |
| | 0.5 | 4% | 0.50 | 0.51 | 0.51 | 0.49 | 3% | 0.50 | 0.51 | 0.51 | 0.49 |
| 0.5 | -0.5 | 8% | 0.26 | 0.19 | 0.19 | 0.24 | 6% | 0.50 | 0.50 | 0.51 | 0.49 |
| | 0 | 8% | 0.31 | 0.25 | 0.24 | 0.31 | 4% | 0.50 | 0.51 | 0.49 | 0.49 |
| | 0.5 | 3% | 0.46 | 0.45 | 0.45 | 0.46 | 5% | 0.50 | 0.50 | 0.49 | 0.50 |
| | | Mixed confounding ($\alpha_2$ =0.8 and $\beta_2$ = -0.8) | | | | | | | | | |
| -0.5 | -0.5 | 4% | 0.52 | 0.53 | 0.53 | 0.52 | 4% | 0.50 | 0.50 | 0.50 | 0.50 |
| | 0 | 6% | 0.48 | 0.48 | 0.46 | 0.47 | 4% | 0.51 | 0.51 | 0.51 | 0.49 |
| | 0.5 | 4% | 0.41 | 0.39 | 0.39 | 0.42 | 3% | 0.50 | 0.50 | 0.50 | 0.50 |
| 0 | -0.5 | 7% | 0.50 | 0.50 | 0.51 | 0.50 | 7% | 0.50 | 0.51 | 0.51 | 0.50 |
| | 0 | 5% | 0.50 | 0.50 | 0.50 | 0.50 | 5% | 0.50 | 0.50 | 0.50 | 0.50 |
| | 0.5 | 5% | 0.50 | 0.50 | 0.49 | 0.50 | 5% | 0.49 | 0.50 | 0.49 | 0.50 |
| 0.5 | -0.5 | 5% | 0.41 | 0.39 | 0.39 | 0.41 | 5% | 0.50 | 0.50 | 0.51 | 0.50 |
| | 0 | 4% | 0.48 | 0.47 | 0.47 | 0.48 | 4% | 0.50 | 0.50 | 0.50 | 0.50 |
| | 0.5 | 6% | 0.52 | 0.54 | 0.52 | 0.52 | 6% | 0.49 | 0.51 | 0.50 | 0.50 |

Proportion homogeneity rejected represents the proportion of simulated datasets where the null hypothesis of homogeneity is rejected

**Supplementary Table 5.** Scenario A2, results for positive, negative, and mixed confounding, stratification approach, $\alpha_1$=0.05

| | | Positive confounding ($\alpha_2$ and $\beta_2$ = 0.8) | | | | | | | | | |
|---|---|---|---|---|---|---|---|---|---|---|---|
| | | Stratifying on collider, C | | | | | Stratifying on residual collider, $C_0$ | | | | |
| $\mu_1$ | $\mu_2$ | Proportion homogeneity rejected (%) | Median estimates Q1 | Median estimates Q2 | Median estimates Q3 | Median estimates Q4 | Proportion homogeneity rejected (%) | Median estimates Q1 | Median estimates Q2 | Median estimates Q3 | Median estimates Q4 |
| -0.5 | -0.5 | 7% | 0.29 | 0.21 | 0.19 | 0.25 | 6% | 0.53 | 0.51 | 0.48 | 0.50 |
| | 0 | 6% | 0.24 | 0.26 | 0.23 | 0.32 | 5% | 0.45 | 0.50 | 0.48 | 0.51 |
| | 0.5 | 6% | 0.37 | 0.48 | 0.39 | 0.41 | 5% | 0.45 | 0.56 | 0.46 | 0.47 |
| 0 | -0.5 | 6% | 0.48 | 0.48 | 0.53 | 0.51 | 6% | 0.49 | 0.51 | 0.49 | 0.52 |
| | 0 | 4% | 0.49 | 0.47 | 0.54 | 0.47 | 4% | 0.49 | 0.49 | 0.55 | 0.47 |
| | 0.5 | 5% | 0.57 | 0.49 | 0.53 | 0.49 | 5% | 0.55 | 0.51 | 0.58 | 0.48 |
| 0.5 | -0.5 | 8% | 0.44 | 0.50 | 0.45 | 0.48 | 8% | 0.50 | 0.53 | 0.47 | 0.54 |
| | 0 | 6% | 0.31 | 0.20 | 0.27 | 0.31 | 6% | 0.52 | 0.44 | 0.54 | 0.53 |
| | 0.5 | 6% | 0.24 | 0.16 | 0.16 | 0.23 | 5% | 0.49 | 0.51 | 0.50 | 0.48 |
| | | Negative confounding ($\alpha_2$ and $\beta_2$ = -0.8) | | | | | | | | | |
| -0.5 | -0.5 | 5% | 0.49 | 0.43 | 0.45 | 0.48 | 5% | 0.50 | 0.45 | 0.47 | 0.48 |
| | 0 | 5% | 0.28 | 0.21 | 0.25 | 0.28 | 5% | 0.46 | 0.50 | 0.53 | 0.47 |
| | 0.5 | 4% | 0.30 | 0.22 | 0.14 | 0.29 | 4% | 0.54 | 0.52 | 0.49 | 0.56 |
| 0 | -0.5 | 4% | 0.49 | 0.52 | 0.48 | 0.52 | 5% | 0.49 | 0.55 | 0.47 | 0.54 |
| | 0 | 4% | 0.55 | 0.46 | 0.46 | 0.48 | 4% | 0.53 | 0.48 | 0.49 | 0.46 |
| | 0.5 | 6% | 0.48 | 0.49 | 0.47 | 0.57 | 6% | 0.47 | 0.50 | 0.49 | 0.58 |
| 0.5 | -0.5 | 7% | 0.26 | 0.22 | 0.19 | 0.24 | 6% | 0.51 | 0.48 | 0.52 | 0.49 |
| | 0 | 4% | 0.32 | 0.23 | 0.33 | 0.33 | 5% | 0.52 | 0.51 | 0.59 | 0.52 |
| | 0.5 | 6% | 0.49 | 0.40 | 0.40 | 0.46 | 6% | 0.52 | 0.50 | 0.45 | 0.47 |
| | | Mixed confounding ($\alpha_2$ = 0.8 and $\beta_2$ = -0.8) | | | | | | | | | |
| -0.5 | -0.5 | 6% | 0.51 | 0.52 | 0.55 | 0.49 | 5% | 0.48 | 0.49 | 0.51 | 0.49 |
| | 0 | 6% | 0.47 | 0.46 | 0.41 | 0.50 | 6% | 0.50 | 0.51 | 0.46 | 0.52 |
| | 0.5 | 6% | 0.44 | 0.39 | 0.35 | 0.38 | 6% | 0.54 | 0.51 | 0.51 | 0.46 |
| 0 | -0.5 | 6% | 0.50 | 0.52 | 0.49 | 0.53 | 6% | 0.52 | 0.50 | 0.50 | 0.53 |
| | 0 | 6% | 0.50 | 0.55 | 0.53 | 0.49 | 5% | 0.48 | 0.55 | 0.53 | 0.50 |
| | 0.5 | 6% | 0.51 | 0.51 | 0.45 | 0.51 | 6% | 0.53 | 0.50 | 0.47 | 0.54 |
| 0.5 | -0.5 | 5% | 0.45 | 0.41 | 0.32 | 0.42 | 6% | 0.53 | 0.53 | 0.43 | 0.50 |
| | 0 | 5% | 0.49 | 0.46 | 0.45 | 0.45 | 5% | 0.51 | 0.52 | 0.46 | 0.46 |
| | 0.5 | 5% | 0.49 | 0.54 | 0.50 | 0.54 | 4% | 0.49 | 0.48 | 0.49 | 0.52 |

Proportion homogeneity rejected represents the proportion of simulated datasets where the null hypothesis of homogeneity is rejected

**Supplementary Table 6.** Scenario A2, results for positive confounding, stratification approach, when $\alpha_1$=0.5 and n=50,000, respectively

| | | Positive confounding ($\alpha_2$ and $\beta_2$ = 0.8), $\alpha_1$=0.5, n =10,000 | | | | | | | | | |
|---|---|---|---|---|---|---|---|---|---|---|---|
| | | Stratifying on collider, C | | | | | Stratifying on residual collider, $C_0$ | | | | |
| $\mu_1$ | $\mu_2$ | Proportion homogeneity rejected (%) | Median estimates Q1 | Median estimates Q2 | Median estimates Q3 | Median estimates Q4 | Proportion homogeneity rejected (%) | Median estimates Q1 | Median estimates Q2 | Median estimates Q3 | Median estimates Q4 |
| -0.5 | -0.5 | 16% | 0.26 | 0.19 | 0.19 | 0.26 | 6% | 0.5 | 0.5 | 0.5 | 0.5 |
| | 0 | 10% | 0.31 | 0.25 | 0.25 | 0.31 | 5% | 0.5 | 0.5 | 0.5 | 0.5 |
| | 0.5 | 5% | 0.46 | 0.45 | 0.45 | 0.46 | 4% | 0.5 | 0.5 | 0.5 | 0.5 |
| 0 | -0.5 | 5% | 0.5 | 0.5 | 0.5 | 0.5 | 5% | 0.5 | 0.5 | 0.5 | 0.5 |
| | 0 | 6% | 0.5 | 0.5 | 0.5 | 0.5 | 5% | 0.49 | 0.5 | 0.5 | 0.5 |
| | 0.5 | 4% | 0.5 | 0.51 | 0.5 | 0.5 | 6% | 0.5 | 0.51 | 0.5 | 0.5 |
| 0.5 | -0.5 | 4% | 0.46 | 0.45 | 0.45 | 0.46 | 4% | 0.5 | 0.5 | 0.5 | 0.5 |
| | 0 | 9% | 0.31 | 0.25 | 0.25 | 0.31 | 6% | 0.5 | 0.49 | 0.5 | 0.5 |
| | 0.5 | 14% | 0.26 | 0.18 | 0.19 | 0.26 | 5% | 0.5 | 0.5 | 0.5 | 0.5 |
| | | Positive confounding ($\alpha_2$ and $\beta_2$ = 0.8), $\alpha_1$=0.1, n =50,000 | | | | | | | | | |
| -1 | -1 | 11% | 0.12 | 0 | 0.01 | 0.13 | 6% | 0.51 | 0.49 | 0.49 | 0.53 |
| | 0 | 16% | 0.09 | -0.04 | -0.02 | 0.09 | 5% | 0.51 | 0.51 | 0.5 | 0.49 |
| | 1 | 4% | 0.36 | 0.4 | 0.38 | 0.39 | 3% | 0.49 | 0.48 | 0.53 | 0.5 |
| -0.5 | -1 | 9% | 0.27 | 0.21 | 0.22 | 0.27 | 4% | 0.47 | 0.51 | 0.51 | 0.5 |
| | 0 | 7% | 0.31 | 0.23 | 0.26 | 0.32 | 5% | 0.5 | 0.49 | 0.5 | 0.52 |
| | 1 | 4% | 0.59 | 0.63 | 0.61 | 0.62 | 2% | 0.48 | 0.48 | 0.47 | 0.49 |
| 0 | -1 | 7% | 0.5 | 0.49 | 0.49 | 0.5 | 10% | 0.49 | 0.49 | 0.51 | 0.5 |
| | 0 | 6% | 0.48 | 0.52 | 0.53 | 0.51 | 6% | 0.47 | 0.52 | 0.52 | 0.51 |
| | 1 | 4% | 0.48 | 0.53 | 0.5 | 0.5 | 3% | 0.48 | 0.54 | 0.49 | 0.51 |
| 0.5 | -1 | 4% | 0.61 | 0.65 | 0.65 | 0.63 | 7% | 0.5 | 0.48 | 0.52 | 0.5 |
| | 0 | 7% | 0.31 | 0.29 | 0.23 | 0.29 | 4% | 0.51 | 0.52 | 0.48 | 0.48 |
| | 1 | 6% | 0.27 | 0.21 | 0.21 | 0.28 | 5% | 0.48 | 0.51 | 0.5 | 0.49 |
| 1 | -1 | 5% | 0.4 | 0.37 | 0.37 | 0.39 | 7% | 0.5 | 0.49 | 0.49 | 0.49 |
| | 0 | 10% | 0.08 | -0.04 | -0.03 | 0.06 | 6% | 0.49 | 0.49 | 0.49 | 0.5 |
| | 1 | 13% | 0.12 | 0 | 0.01 | 0.11 | 4% | 0.52 | 0.5 | 0.51 | 0.49 |

Proportion homogeneity rejected represents the proportion of simulated datasets where the null hypothesis of homogeneity is rejected

**Supplementary Table 7.** Scenario A3, results for negative, and mixed confounding, stratification approach, $\alpha_1$=0.1

| | | Negative confounding ($\alpha_2$ and $\beta_2$ = -0.8) | | | | | | | | | |
|---|---|---|---|---|---|---|---|---|---|---|---|
| | | Stratifying on collider, C | | | | | Stratifying on residual collider, $C_0$ | | | | |
| $\mu_1$ | $\mu_2$ | Proportion homogeneity rejected (%) | Median estimates Q1 | Median estimates Q2 | Median estimates Q3 | Median estimates Q4 | Proportion homogeneity rejected (%) | Median estimates Q1 | Median estimates Q2 | Median estimates Q3 | Median estimates Q4 |
| -0.5 | -0.5 | 15% | 0.22 | 0.45 | 0.55 | 0.76 | 21% | 0.22 | 0.45 | 0.55 | 0.76 |
| | 0 | 19% | 0.10 | 0.23 | 0.33 | 0.61 | 29% | 0.10 | 0.23 | 0.33 | 0.61 |
| | 0.5 | 24% | -0.03 | 0.14 | 0.30 | 0.64 | 50% | -0.03 | 0.14 | 0.30 | 0.64 |
| 0 | -0.5 | 17% | 0.31 | 0.53 | 0.68 | 0.87 | 16% | 0.31 | 0.53 | 0.68 | 0.87 |
| | 0 | 14% | 0.34 | 0.54 | 0.61 | 0.88 | 13% | 0.34 | 0.54 | 0.61 | 0.88 |
| | 0.5 | 15% | 0.28 | 0.54 | 0.66 | 0.93 | 15% | 0.28 | 0.54 | 0.66 | 0.93 |
| 0.5 | -0.5 | 23% | 0.10 | 0.23 | 0.36 | 0.72 | 42% | 0.10 | 0.23 | 0.36 | 0.72 |
| | 0 | 13% | 0.18 | 0.27 | 0.43 | 0.69 | 29% | 0.18 | 0.27 | 0.43 | 0.69 |
| | 0.5 | 14% | 0.32 | 0.47 | 0.66 | 0.89 | 22% | 0.32 | 0.47 | 0.66 | 0.89 |
| | | Mixed confounding ($\alpha_2$ = 0.8 and $\beta_2$ = -0.8) | | | | | | | | | |
| -0.5 | -0.5 | 32% | 0.22 | 0.45 | 0.55 | 0.76 | 63% | 0.02 | 0.40 | 0.60 | 1.01 |
| | 0 | 18% | 0.10 | 0.23 | 0.33 | 0.61 | 43% | 0.14 | 0.40 | 0.62 | 0.89 |
| | 0.5 | 23% | -0.03 | 0.14 | 0.30 | 0.64 | 35% | 0.15 | 0.42 | 0.61 | 0.84 |
| 0 | -0.5 | 27% | 0.31 | 0.53 | 0.68 | 0.87 | 27% | 0.29 | 0.55 | 0.67 | 0.90 |
| | 0 | 23% | 0.34 | 0.54 | 0.61 | 0.88 | 24% | 0.35 | 0.54 | 0.67 | 0.89 |
| | 0.5 | 27% | 0.28 | 0.54 | 0.66 | 0.93 | 27% | 0.34 | 0.53 | 0.68 | 0.91 |
| 0.5 | -0.5 | 35% | 0.10 | 0.23 | 0.36 | 0.72 | 36% | 0.38 | 0.62 | 0.79 | 1.03 |
| | 0 | 20% | 0.18 | 0.27 | 0.43 | 0.69 | 43% | 0.30 | 0.60 | 0.80 | 1.10 |
| | 0.5 | 26% | 0.32 | 0.47 | 0.66 | 0.89 | 56% | 0.25 | 0.56 | 0.81 | 1.17 |

Proportion homogeneity rejected represents the proportion of simulated datasets where the null hypothesis of homogeneity is rejected

**Supplementary Table 8.** Scenario A3, results for positive, negative, and mixed confounding, stratification approach, $\alpha_1$=0.3

| | | Positive confounding ($\alpha_2$ and $\beta_2$ = 0.8) | | | | | | | | | |
|---|---|---|---|---|---|---|---|---|---|---|---|
| | | Stratifying on collider, C | | | | | Stratifying on residual collider, $C_0$ | | | | |
| $\mu_1$ | $\mu_2$ | Proportion homogeneity rejected (%) | Median estimates Q1 | Median estimates Q2 | Median estimates Q3 | Median estimates Q4 | Proportion homogeneity rejected (%) | Median estimates Q1 | Median estimates Q2 | Median estimates Q3 | Median estimates Q4 |
| -0.5 | -0.5 | 98% | -0.04 | 0.15 | 0.29 | 0.61 | 100% | 0.04 | 0.37 | 0.62 | 0.99 |
| | 0 | 88% | 0.06 | 0.22 | 0.34 | 0.61 | 100% | 0.10 | 0.40 | 0.61 | 0.88 |
| | 0.5 | 81% | 0.24 | 0.42 | 0.55 | 0.76 | 96% | 0.18 | 0.42 | 0.58 | 0.84 |
| 0 | -0.5 | 88% | 0.31 | 0.54 | 0.67 | 0.88 | 88% | 0.31 | 0.54 | 0.67 | 0.88 |
| | 0 | 76% | 0.35 | 0.55 | 0.65 | 0.85 | 76% | 0.34 | 0.55 | 0.66 | 0.85 |
| | 0.5 | 90% | 0.31 | 0.53 | 0.67 | 0.88 | 89% | 0.31 | 0.53 | 0.68 | 0.88 |
| 0.5 | -0.5 | 75% | 0.34 | 0.52 | 0.64 | 0.87 | 93% | 0.37 | 0.61 | 0.78 | 1.03 |
| | 0 | 82% | 0.17 | 0.30 | 0.43 | 0.70 | 99% | 0.31 | 0.60 | 0.80 | 1.10 |
| | 0.5 | 98% | 0.07 | 0.23 | 0.37 | 0.70 | 100% | 0.23 | 0.58 | 0.82 | 1.17 |
| | | Negative confounding ($\alpha_2$ and $\beta_2$ = -0.8) | | | | | | | | | |
| -0.5 | -0.5 | 85% | 0.23 | 0.43 | 0.55 | 0.79 | 97% | 0.16 | 0.42 | 0.59 | 0.84 |
| | 0 | 86% | 0.06 | 0.23 | 0.33 | 0.60 | 100% | 0.11 | 0.40 | 0.59 | 0.88 |
| | 0.5 | 98% | -0.04 | 0.15 | 0.29 | 0.60 | 100% | 0.02 | 0.38 | 0.62 | 0.97 |
| 0 | -0.5 | 88% | 0.31 | 0.53 | 0.66 | 0.88 | 88% | 0.31 | 0.53 | 0.66 | 0.88 |
| | 0 | 76% | 0.35 | 0.53 | 0.67 | 0.85 | 77% | 0.35 | 0.53 | 0.66 | 0.85 |
| | 0.5 | 89% | 0.31 | 0.52 | 0.67 | 0.89 | 88% | 0.31 | 0.53 | 0.67 | 0.89 |
| 0.5 | -0.5 | 97% | 0.06 | 0.24 | 0.36 | 0.70 | 100% | 0.23 | 0.58 | 0.82 | 1.16 |
| | 0 | 83% | 0.16 | 0.30 | 0.42 | 0.71 | 99% | 0.31 | 0.59 | 0.80 | 1.09 |
| | 0.5 | 77% | 0.34 | 0.51 | 0.65 | 0.88 | 93% | 0.37 | 0.61 | 0.79 | 1.03 |
| | | Mixed confounding ($\alpha_2$ = 0.8 and $\beta_2$ = -0.8) | | | | | | | | | |
| -0.5 | -0.5 | 100% | 0.23 | 0.50 | 0.63 | 0.87 | 100% | 0.04 | 0.38 | 0.62 | 0.97 |
| | 0 | 99% | 0.23 | 0.45 | 0.55 | 0.78 | 100% | 0.10 | 0.40 | 0.60 | 0.89 |
| | 0.5 | 99% | 0.18 | 0.37 | 0.50 | 0.72 | 100% | 0.17 | 0.42 | 0.59 | 0.84 |
| 0 | -0.5 | 100% | 0.31 | 0.52 | 0.67 | 0.89 | 100% | 0.31 | 0.52 | 0.66 | 0.89 |
| | 0 | 98% | 0.35 | 0.53 | 0.66 | 0.86 | 98% | 0.35 | 0.53 | 0.66 | 0.86 |
| | 0.5 | 99% | 0.31 | 0.53 | 0.68 | 0.88 | 99% | 0.31 | 0.53 | 0.68 | 0.88 |
| 0.5 | -0.5 | 98% | 0.30 | 0.46 | 0.58 | 0.83 | 100% | 0.38 | 0.62 | 0.78 | 1.04 |
| | 0 | 98% | 0.33 | 0.52 | 0.64 | 0.88 | 100% | 0.31 | 0.60 | 0.80 | 1.09 |
| | 0.5 | 100% | 0.32 | 0.56 | 0.71 | 0.97 | 100% | 0.23 | 0.58 | 0.82 | 1.17 |

Proportion homogeneity rejected represents the proportion of simulated datasets where the null hypothesis of homogeneity is rejected

**Supplementary Table 9.** Scenario A3, results for positive, negative, and mixed confounding, stratification approach, $\alpha_1$=0.05

| | | **Positive confounding ($\alpha_2$ and $\beta_2$ = 0.8)** | | | | | | | | | |
|---|---|---|---|---|---|---|---|---|---|---|---|
| | | **Stratifying on collider, C** | | | | | **Stratifying on residual collider, $C_0$** | | | | |
| **$\mu_1$** | **$\mu_2$** | **Proportion homogeneity rejected (%)** | **Median estimates Q1** | **Median estimates Q2** | **Median estimates Q3** | **Median estimates Q4** | **Proportion homogeneity rejected (%)** | **Median estimates Q1** | **Median estimates Q2** | **Median estimates Q3** | **Median estimates Q4** |
| -0.5 | -0.5 | 10% | -0.02 | 0.14 | 0.32 | 0.63 | 17% | 0.05 | 0.35 | 0.66 | 0.99 |
| | 0 | 8% | 0.11 | 0.22 | 0.31 | 0.53 | 9% | 0.14 | 0.38 | 0.60 | 0.85 |
| | 0.5 | 8% | 0.22 | 0.44 | 0.58 | 0.77 | 9% | 0.17 | 0.41 | 0.61 | 0.86 |
| 0 | -0.5 | 8% | 0.33 | 0.57 | 0.66 | 0.79 | 8% | 0.33 | 0.55 | 0.66 | 0.81 |
| | 0 | 6% | 0.27 | 0.48 | 0.65 | 0.84 | 6% | 0.27 | 0.51 | 0.64 | 0.86 |
| | 0.5 | 7% | 0.32 | 0.50 | 0.73 | 0.93 | 8% | 0.33 | 0.51 | 0.75 | 0.93 |
| 0.5 | -0.5 | 4% | 0.37 | 0.48 | 0.66 | 0.89 | 7% | 0.41 | 0.61 | 0.85 | 1.03 |
| | 0 | 6% | 0.15 | 0.34 | 0.41 | 0.68 | 9% | 0.27 | 0.60 | 0.79 | 1.05 |
| | 0.5 | 8% | 0.03 | 0.26 | 0.41 | 0.71 | 16% | 0.18 | 0.60 | 0.84 | 1.19 |
| | | **Negative confounding ($\alpha_2$ and $\beta_2$ = -0.8)** | | | | | | | | | |
| -0.5 | -0.5 | 8% | 0.20 | 0.39 | 0.57 | 0.78 | 11% | 0.14 | 0.38 | 0.61 | 0.88 |
| | 0 | 7% | 0.10 | 0.17 | 0.37 | 0.56 | 10% | 0.13 | 0.33 | 0.61 | 0.85 |
| | 0.5 | 7% | 0.01 | 0.14 | 0.27 | 0.60 | 14% | 0.07 | 0.37 | 0.61 | 0.95 |
| 0 | -0.5 | 6% | 0.28 | 0.55 | 0.67 | 0.89 | 8% | 0.26 | 0.56 | 0.69 | 0.88 |
| | 0 | 7% | 0.33 | 0.55 | 0.66 | 0.87 | 6% | 0.35 | 0.55 | 0.65 | 0.90 |
| | 0.5 | 5% | 0.27 | 0.56 | 0.68 | 0.87 | 5% | 0.26 | 0.51 | 0.70 | 0.87 |
| 0.5 | -0.5 | 8% | 0.08 | 0.22 | 0.37 | 0.74 | 14% | 0.26 | 0.55 | 0.80 | 1.19 |
| | 0 | 8% | 0.16 | 0.29 | 0.45 | 0.67 | 10% | 0.31 | 0.61 | 0.81 | 1.07 |
| | 0.5 | 6% | 0.37 | 0.55 | 0.69 | 0.91 | 5% | 0.40 | 0.66 | 0.82 | 1.08 |
| | | **Mixed confounding ($\alpha_2$ = 0.8 and $\beta_2$= -0.8)** | | | | | | | | | |
| -0.5 | -0.5 | 10% | 0.25 | 0.50 | 0.64 | 0.87 | 20% | 0.07 | 0.36 | 0.62 | 0.95 |
| | 0 | 7% | 0.30 | 0.46 | 0.53 | 0.78 | 10% | 0.16 | 0.42 | 0.58 | 0.91 |
| | 0.5 | 8% | 0.18 | 0.35 | 0.53 | 0.73 | 13% | 0.15 | 0.43 | 0.63 | 0.84 |
| 0 | -0.5 | 7% | 0.35 | 0.54 | 0.68 | 0.86 | 8% | 0.35 | 0.54 | 0.67 | 0.88 |
| | 0 | 7% | 0.33 | 0.52 | 0.63 | 0.89 | 8% | 0.34 | 0.53 | 0.63 | 0.90 |
| | 0.5 | 9% | 0.31 | 0.57 | 0.66 | 0.88 | 9% | 0.30 | 0.58 | 0.65 | 0.88 |
| 0.5 | -0.5 | 8% | 0.29 | 0.42 | 0.54 | 0.77 | 9% | 0.39 | 0.58 | 0.74 | 0.99 |
| | 0 | 9% | 0.31 | 0.47 | 0.61 | 0.87 | 14% | 0.26 | 0.55 | 0.78 | 1.09 |
| | 0.5 | 11% | 0.31 | 0.55 | 0.75 | 0.99 | 20% | 0.20 | 0.55 | 0.85 | 1.19 |

Proportion homogeneity rejected represents the proportion of simulated datasets where the null hypothesis of homogeneity is rejected

**Supplementary Table 10.** Scenario B1, results for positive confounding, adjustment approach, $\alpha_1$=0.1, considering C as a function of both risk factor and outcome

| | | Positive confounding ($\alpha_2$ and $\beta_2$= 0.8) | | | |
|---|---|---|---|---|---|
| | | **Median estimate** | **Type I error rate (%)** | **Median estimate** | **Type I error rate (%)** |
| **$\mu_1$** | **$\mu_3$** | *No adjust for collider* | | *Adjust Y/G for collider* | |
| -0.5 | -0.5 | -0.01 | 4% | -0.27 | 65% |
| | 0 | 0.00 | 7% | -0.03 | 7% |
| | 0.5 | -0.01 | 5% | 0.23 | 52% |
| 0 | -0.5 | 0.01 | 6% | 0.00 | 5% |
| | 0 | 0.00 | 6% | 0.00 | 6% |
| | 0.5 | 0.00 | 5% | 0.01 | 7% |
| 0.5 | -0.5 | -0.01 | 6% | 0.08 | 8% |
| | 0 | -0.01 | 3% | -0.17 | 28% |
| | 0.5 | 0.02 | 5% | -0.25 | 74% |

Empirical Type I error rate represents the proportion of simulated datasets where the null hypothesis is not rejected

**Supplementary Table 11.** Scenario B2 and B3, results for positive confounding, stratification approach, $\alpha_1$=0.1, considering C as a function of both risk factor and outcome, when the causal effect is constant (Scenario B2) and it when depends on U (Scenario B3)

| | | Scenario B2 | | | Positive confounding ($\alpha_2$ and $\beta_2$= 0.8), where $\beta_1$=0.5 | | | | | | |
|---|---|---|---|---|---|---|---|---|---|---|---|
| | | Stratifying on collider, C | | | | | Stratifying on residual collider, $C_0$ | | | | |
| $\mu_1$ | $\mu_3$ | Proportion homogeneity rejected (%) | Median estimates Q1 | Median estimates Q2 | Median estimates Q3 | Median estimates Q4 | Proportion homogeneity rejected (%) | Median estimates Q1 | Median estimates Q2 | Median estimates Q3 | Median estimates Q4 |
| -0.5 | -0.5 | 8% | 0.07 | -0.05 | -0.03 | 0.06 | 6% | 0.49 | 0.49 | 0.50 | 0.51 |
| | 0 | 5% | 0.39 | 0.37 | 0.37 | 0.37 | 5% | 0.51 | 0.49 | 0.52 | 0.51 |
| | 0.5 | 5% | 0.60 | 0.66 | 0.65 | 0.61 | 5% | 0.47 | 0.52 | 0.49 | 0.49 |
| 0 | -0.5 | 6% | 0.36 | 0.32 | 0.35 | 0.34 | 6% | 0.50 | 0.51 | 0.51 | 0.49 |
| | 0 | 7% | 0.48 | 0.50 | 0.52 | 0.48 | 6% | 0.49 | 0.52 | 0.51 | 0.48 |
| | 0.5 | 5% | 0.35 | 0.31 | 0.32 | 0.36 | 5% | 0.50 | 0.50 | 0.51 | 0.50 |
| 0.5 | -0.5 | 5% | 0.54 | 0.54 | 0.58 | 0.54 | 5% | 0.49 | 0.49 | 0.53 | 0.50 |
| | 0 | 6% | 0.24 | 0.21 | 0.20 | 0.28 | 5% | 0.48 | 0.52 | 0.49 | 0.52 |
| | 0.5 | 6% | 0.10 | 0.01 | 0.02 | 0.10 | 5% | 0.48 | 0.51 | 0.51 | 0.50 |
| | | Scenario B3 | | | Positive confounding ($\alpha_2$ and $\beta_2$= 0.8), where $\beta_1$=0.5+0.2U | | | | | | |
| $\mu_1$ | $\mu_3$ | Proportion homogeneity rejected (%) | Median estimates Q1 | Median estimates Q2 | Median estimates Q3 | Median estimates Q4 | Proportion homogeneity rejected (%) | Median estimates Q1 | Median estimates Q2 | Median estimates Q3 | Median estimates Q4 |
| -0.5 | -0.5 | 6% | 0.09 | -0.07 | -0.07 | 0.00 | 9% | 0.65 | 0.52 | 0.45 | 0.35 |
| | 0 | 7% | 0.40 | 0.35 | 0.38 | 0.38 | 7% | 0.52 | 0.50 | 0.51 | 0.48 |
| | 0.5 | 12% | 0.45 | 0.59 | 0.69 | 0.85 | 11% | 0.32 | 0.45 | 0.52 | 0.71 |
| 0 | -0.5 | 7% | 0.50 | 0.32 | 0.27 | 0.24 | 8% | 0.69 | 0.50 | 0.43 | 0.36 |
| | 0 | 5% | 0.43 | 0.46 | 0.51 | 0.57 | 4% | 0.42 | 0.48 | 0.50 | 0.56 |
| | 0.5 | 9% | 0.20 | 0.28 | 0.32 | 0.51 | 13% | 0.34 | 0.47 | 0.53 | 0.70 |
| 0.5 | -0.5 | 7% | 0.66 | 0.58 | 0.58 | 0.41 | 7% | 0.63 | 0.52 | 0.48 | 0.36 |
| | 0 | 5% | 0.17 | 0.19 | 0.21 | 0.34 | 6% | 0.36 | 0.49 | 0.55 | 0.64 |
| | 0.5 | 8% | -0.01 | -0.03 | -0.02 | 0.16 | 13% | 0.31 | 0.46 | 0.53 | 0.69 |

Proportion homogeneity rejected represents the proportion of simulated datasets where the null hypothesis of homogeneity is rejected

**Supplementary Table 12.** Scenario C1, results for positive confounding, adjustment approach, $\alpha_1$=0.1, considering a binary outcome Y

| | | Positive confounding ($\alpha_2$ and $\beta_2$= 0.8) | | | |
|---|---|---|---|---|---|
| | | **Median estimate** | **Type I error rate (%)** | **Median estimate** | **Type I error rate (%)** |
| **$\mu_1$** | **$\mu_2$** | *No adjust for collider* | | *Adjust Y/G for collider* | |
| -0.5 | -0.5 | 0.02 | 6% | -0.14 | 13% |
| | 0 | -0.01 | 5% | -0.11 | 8% |
| | 0.5 | -0.01 | 4% | 0.02 | 5% |
| 0 | -0.5 | -0.01 | 4% | -0.01 | 4% |
| | 0 | -0.01 | 4% | -0.01 | 4% |
| | 0.5 | 0.00 | 5% | 0.00 | 5% |
| 0.5 | -0.5 | 0.00 | 4% | 0.02 | 6% |
| | 0 | 0.02 | 5% | -0.09 | 7% |
| | 0.5 | -0.01 | 6% | -0.18 | 14% |

Empirical Type I error rate represents the proportion of simulated datasets where the null hypothesis is not rejected

**Supplementary Table 13.** Scenario C2 and C3, results for positive confounding, stratification approach, $\alpha_1$=0.1, considering a binary outcome Y, when the causal effect is constant (Scenario C2) and it when depends on C (Scenario C3)

| | | Scenario C2 **Positive confounding ($\alpha_2$ and $\beta_2$= 0.8), where $\beta_1$=0.5** | | | | | | | | | |
|---|---|---|---|---|---|---|---|---|---|---|---|
| | | **Stratifying on collider, C** | | | | | **Stratifying on residual collider, $C_0$** | | | | |
| **$\mu_1$** | **$\mu_2$** | **Proportion homogeneity rejected (%)** | **Median estimates Q1** | **Median estimates Q2** | **Median estimates Q3** | **Median estimates Q4** | **Proportion homogeneity rejected (%)** | **Median estimates Q1** | **Median estimates Q2** | **Median estimates Q3** | **Median estimates Q4** |
| -0.5 | -0.5 | 8% | 0.17 | 0.16 | 0.15 | 0.22 | 6% | 0.43 | 0.42 | 0.39 | 0.41 |
| | 0 | 6% | 0.21 | 0.19 | 0.18 | 0.22 | 7% | 0.37 | 0.38 | 0.40 | 0.36 |
| | 0.5 | 5% | 0.39 | 0.37 | 0.38 | 0.36 | 4% | 0.42 | 0.40 | 0.39 | 0.38 |
| 0 | -0.5 | 4% | 0.44 | 0.41 | 0.37 | 0.37 | 4% | 0.43 | 0.39 | 0.37 | 0.37 |
| | 0 | 3% | 0.41 | 0.42 | 0.41 | 0.38 | 4% | 0.41 | 0.39 | 0.39 | 0.36 |
| | 0.5 | 5% | 0.38 | 0.41 | 0.39 | 0.40 | 4% | 0.40 | 0.41 | 0.39 | 0.40 |
| 0.5 | -0.5 | 8% | 0.28 | 0.29 | 0.36 | 0.38 | 7% | 0.32 | 0.35 | 0.37 | 0.41 |
| | 0 | 6% | 0.23 | 0.20 | 0.19 | 0.23 | 5% | 0.38 | 0.38 | 0.42 | 0.39 |
| | 0.5 | 5% | 0.27 | 0.16 | 0.13 | 0.23 | 5% | 0.50 | 0.44 | 0.41 | 0.46 |
| | | Scenario C3 **Positive confounding ($\alpha_2$ and $\beta_2$= 0.8), where $\beta_1$=0.5+0.2C** | | | | | | | | | |
| **$\mu_1$** | **$\mu_2$** | **Proportion homogeneity rejected (%)** | **Median estimates Q1** | **Median estimates Q2** | **Median estimates Q3** | **Median estimates Q4** | **Proportion homogeneity rejected (%)** | **Median estimates Q1** | **Median estimates Q2** | **Median estimates Q3** | **Median estimates Q4** |
| -0.5 | -0.5 | 7% | -0.07 | 0.14 | 0.30 | 0.44 | 13% | 0.01 | 0.37 | 0.57 | 0.70 |
| | 0 | 7% | 0.04 | 0.18 | 0.26 | 0.44 | 10% | 0.09 | 0.34 | 0.49 | 0.61 |
| | 0.5 | 6% | 0.24 | 0.39 | 0.41 | 0.49 | 8% | 0.19 | 0.37 | 0.47 | 0.54 |
| 0 | -0.5 | 5% | 0.27 | 0.38 | 0.49 | 0.62 | 5% | 0.26 | 0.39 | 0.49 | 0.60 |
| | 0 | 6% | 0.28 | 0.40 | 0.48 | 0.58 | 8% | 0.27 | 0.40 | 0.49 | 0.58 |
| | 0.5 | 6% | 0.31 | 0.39 | 0.52 | 0.61 | 6% | 0.29 | 0.40 | 0.53 | 0.62 |
| 0.5 | -0.5 | 7% | 0.22 | 0.40 | 0.50 | 0.60 | 10% | 0.23 | 0.43 | 0.58 | 0.66 |
| | 0 | 5% | 0.14 | 0.23 | 0.33 | 0.50 | 7% | 0.26 | 0.41 | 0.59 | 0.73 |
| | 0.5 | 6% | 0.06 | 0.21 | 0.27 | 0.42 | 11% | 0.22 | 0.50 | 0.62 | 0.78 |

Proportion homogeneity rejected represents the proportion of simulated datasets where the null hypothesis of homogeneity is rejected
